# Effect of *Andrographis paniculata* treatment for patients with early-stage COVID-19 on the prevention of pneumonia: A retrospective cohort study

**DOI:** 10.1101/2023.03.15.23287287

**Authors:** Amporn Benjaponpitak, Thiti Sawaengtham, Tewan Thaneerat, Kulthanit Wanaratna, Palang Chotsiri, Chalermquan Rungsawang, Sakkarin Bhubhanil, Sataporn Charoensuk, Suwat Benjaponpitak, Sarawut Lapmanee, Sayomporn Sirinavin

## Abstract

There is a need for safe and cost-effective treatments for COVID-19. *Andrographis paniculata* (AP) is an herbal plant that has been used for centuries to treat upper respiratory tract infections. Andrographolide is the major active component of AP that inhibits intracellular SARS-CoV-2 replication and has anti-inflammatory action. We performed a retrospective cohort study to evaluate the therapeutic and adverse effects of oral AP-products on COVID-19 by using the risk of pneumonia diagnosed by chest radiography as an indicator. This study included patients 15 to 60 years of age with laboratory-confirmed early-stage (asymptomatic or mild) COVID-19 without comorbidities at seven hospitals in three adjacent provinces in Thailand, between December 2020 and March 2021. Patients were treated for five days with either AP-extract (60 mg andrographolide, 3 times daily) or crude-AP (48 mg andrographolide, 3 times daily), when available. Patient information was prospectively recorded in the structured medical records and retrospectively reviewed. All eligible patients who received AP-treatment were included and control patients who did not receive AP-treatment were randomly selected using a ratio of approximately 1:1. Pneumonia occurred in 1/243 AP-treatment patients and 69/285 control patients. The risks of pneumonia after adjusting for confounding effects were 0.3% (95%CI, 0%-0.9%) and 24.3% (95%CI, 19.0%-29.7%) in the AP-treatment and control groups, respectively. The number needed to treat to avoid pneumonia development in one patient was four (95% CI, 3-5). Eight patients developed mild adverse events. AP-treatment regimens are acceptably safe and associated with highly reduced rates of pneumonia for patients with early-stage COVID-19.

## 1. Introduction

Coronavirus disease 2019 (COVID-19), which is caused by the novel severe acute respiratory syndrome coronavirus 2 (SARS-CoV-2), was first reported in December 2019, and it resulted in a global pandemic (World Health Organization, 2022a). More cost-effective and widely available interventions to minimise losses caused by COVID-19 are still needed, especially in resource-limited countries. The clinical spectrum of COVID-19 is categorised as asymptomatic or pre-symptomatic, mild, moderate, severe, and critical. Patients with “early-stage COVID-19” are asymptomatic or have mild symptoms that are generally self-limited but could progress. Approximately 2 to 3 days after infection, patients with mild COVID-19 present with a variety of symptoms, including fever, headaches, myalgia, fatigue, nasal congestion, dry cough, sore throat, loss of taste and/or smell, and diarrhoea; however, they do not present with pneumonia. Patients with moderate COVID-19 commonly experience pneumonia development; these patients require hospitalisation or admission to an intensive care facility. Furthermore, some can experience respiratory failure, which may result in chronic lung diseases or death. Older age, male sex, and comorbidities have been associated with worse COVID-19 outcomes (Wu and McGoogan, 2020; Centers for Disease Control and Prevention USA, 2022b; National Institutes of Health USA, 2022). Early during its clinical course, COVID-19 is primarily driven by the intracellular replication of SARS-CoV-2 and upper respiratory tract mucosal cell damage. Later in its clinical course, COVID-19 appears to be driven by lower respiratory tract invasion and dysregulated immune/inflammatory responses to SARS-CoV-2, thus leading to pneumonia and organ tissue damage (Lamers and Haagmans, 2022). Therefore, interventions that are safe, cost-effective, and inhibit viral replication during early-stage COVID-19, thus preventing disease progression, are in high demand.

*Andrographis paniculata* (Burm.f.) Nees (AP) is a eudicot species in the Acanthaceae family (taxonomy ID: NCBI 175694) (National Center for Biotechnology Information, 2022). AP is an annual herb native to the subcontinent of India that has been widely introduced, naturalised, and cultivated in the tropical and subtropical regions of Asia (Kew Science, 2022). This bitter-tasting herb is commonly called green chiretta; however, it has many local names, such as Kalmegh in India, Chuan Xin Lian in China, Sambiloto in Indonesia, Hempedu Bumi in Malaysia, and Fa Thalai Chon in Thailand. AP comprises several components that are known for their anti-microbial, anti-inflammatory, immunomodulatory, anti-oxidant, and vasodilatory effects (Hossain et al., 2021; Zeng et al., 2022). Oral AP has been used for centuries as a traditional medicine in India, China, Thailand, and other Asian countries. It is widely known as a treatment for infectious diseases, fever, common cold, and influenza (National Drug Committee, Thailand, 2000; the State Pharmacopoeia Commission of the People’s Republic of China, 2015; Banerjee et al., 2021). Several double-blind studies have demonstrated the benefits of AP for viral respiratory tract infections (Coon, 2004; Poolsup, 2004). AP is included in The National List of Essential Herbal Drugs A.D. 1999 of Thailand as an herbal drug used for the treatment of common cold symptoms and non-infectious diarrhoea. Andrographolide, which is a bitter bioactive diterpenoid, is the major active component of AP.

*In silico* or computer-based modelling studies have revealed that andrographolide from AP inhibits the main protease of SARS-CoV-2, which is an essential enzyme for viral replication, by successfully docking in its binding site (Shi et al., 2020; Sukardiman et al., 2020; Enmozhi et al., 2021; Hiremath et al., 2021; Murugan et al., 2021). An *in vitro* study of human lung epithelial cells demonstrated that AP extract and andrographolide treatment of cells infected with SARS-CoV-2 inhibited intracellular viral replication (Sa-Ngiamsuntorn et al., 2021). Furthermore, *in silico* and *in vitro* studies indicated that the anti-inflammatory effects of AP bioactive ingredients were related to reductions in the expression of interleukin-6 and tumour necrosis factor-alpha, which regulate inflammation and the immune response (Zhu et al., 2021).

To optimise the dose, the pharmacokinetic parameters of the active components of AP were studied after multiple oral doses were administered to healthy Thai volunteers (Pholphana et al., 2016). Dosage regimens to treat COVID-19 were proposed based on the results of clinical studies published in scientific literature and the anti-SARS-CoV-2 activity of AP extract in tissue culture (Phumiamorn et al., 2020). Preliminary clinical experience with a small group of Thai patients suggested that AP treatment regimens could be effective for mild COVID-19 (Yearsley, 2021).

Two AP products were used as an emergency treatment for early-stage COVID-19 during the second wave of the COVID-19 epidemic in Thailand. We retrospectively evaluated the therapeutic and adverse effects of oral AP products on early-stage COVID-19.

## 2. Methods

### 2.1 Study design

We performed this retrospective cohort study using data collected from patients with asymptomatic or mild COVID-19 admitted in hospitals for isolation and treatment, during the second epidemic wave in Thailand. The information was prospectively recorded in structured medical records as routine practice. The patients received AP treatment when it was available. We compared the risks of pneumonia development for all eligible AP treatment group and randomly chosen control group patients.

### 2.2 Study setting

The second wave of the COVID-19 epidemic in Thailand started in mid-December 2020 in fresh markets in Samut Sakhon Province, which is in the central region of Thailand, and it spread to the adjacent provinces. Management of the spread of SARS-CoV-2 began at the onset of the outbreak. Molecular testing of the nasopharyngeal swab using the real-time reverse-transcriptase polymerase chain reaction to determine the presence of SARS-CoV-2 was performed to find cases in the community and patients who presented to the hospitals. Patients with laboratory-confirmed SARS-CoV-2 were promptly admitted to local hospitals, which were connected as networked medical care facilities, for isolation and treatment. Chest radiography was routinely performed at the time of admission.

All patients with asymptomatic or mild COVID-19 received general supportive care, including rest, hydration, vital sign monitoring, oxygen saturation monitoring, symptom treatment, and measures to prevent virus spread. Patients with more severe disease were treated separately. Patient information at the time of admission and during follow-up were regularly recorded in the structured medical records of COVID-19 patients by medical and paramedical personnel. Those who recovered were discharged from the hospital by day 14 of hospitalisation and followed up for at least 10 days. Those whose condition worsened were moved to medical care facilities for patients with more severe cases. The diagnoses of all patients at the time of discharge were recorded in the structured electronic medical records.

### 2.3 Study drug regimens

The active component of the study drug was andrographolide, which was obtained from AP. During the study period, two AP products were occasionally available as COVID-19 treatments in the included provinces. These were orally administered as capsules containing a standardised ethanol extract of the aerial part of AP (20 mg of andrographolide per capsule; AP extract) or standardised crude AP powder containing 400 mg of dried AP powder with 3% andrographolide (12 mg of andrographolide per capsule; crude AP). The AP extract was produced by Thai Herbal Products Co., Ltd. (Ayutthaya, Thailand), and crude AP was produced by the Chao Phya Abhaibhubejhr Hospital Foundation (Prachin Buri, Thailand). Both are herbal products registered with the Thailand Food and Drug Administration (registration numbers G169/62 and G512/60, respectively). The andrographolide concentration of each AP product was determined using high-performance liquid chromatography. During the 5-day course of treatment, each patient received either AP extract (60 mg of andrographolide, 3 times daily) or crude AP (48 mg of andrographolide, 3 times daily).

During the study period, the only available anti-viral drug was favipiravir; however, it was available in very limited quantities for patients with moderate and severe COVID-19. The AP treatment regimens were initiated after reviewing published information in scientific literature together with more recent information (Pholphana et al., 2016; Phumiamorn et al., 2020; Sa-Ngiamsuntorn et al., 2021; Yearsley, 2021). Shortages of these AP products were common; therefore, each hospital could use them only when they were available. Treatment with an AP product was offered to the patients on the admission date. The opportunity to receive AP treatment was dependent on the availability of AP products.

### 2.4 Patients

The patient inclusion criteria were as follows: age 15 to 60 years; laboratory-confirmed SARS-CoV-2 diagnosed by the real-time reverse-transcriptase polymerase chain reaction; asymptomatic or [mild COVID-19](https://www.healthline.com/health/coronavirus-symptoms) on admission with no evidence of lower respiratory tract problems; and admission to one of seven hospitals for COVID-19 isolation and treatment in three adjacent provinces (Samut Sakhon, Ratchaburi, and Nakhon Pathom) between December 2020 and March 2021. We excluded those with underlying medical conditions associated with a high risk of severe COVID-19, pregnant women, and those who received any anti-viral drugs, herbs, or products containing andrographolide before hospital admission. Although mild obesity (body mass index, 30-34.9 kg/m^2^) has been classified as a risk factor for severe COVID-19, patients with mild obesity were included in this study.

### 2.5 Data collection

Retrospective data collection was conducted from April to May 2021. The AP treatment was identified by reviewing the pharmacy records. All eligible patients treated with the AP regimens were included in the study. The control group included eligible patients who did not receive AP treatment (ratio of 1:1 with the AP treatment group). The medical records of the control patients were randomly and blindly selected from a database of hospitalised COVID-19 patients by non-clinical staff without knowledge of the study protocol and clinical course or outcomes of the patients. A team of physicians reviewed and collected demographic, clinical, and outcome data from the structured medical records of the eligible COVID-19 patients. Patient data were anonymised, and informed consent was not required.

### 2.6 Outcomes

The outcomes of interest were the occurrence of pneumonia confirmed by chest radiography and any probable adverse reactions to AP treatment described in the structured medical records. According to the standard of care of the hospitals during the study period, all patients underwent chest radiography at the time of hospital admission, when they developed suspicious signs or symptoms of lower respiratory tract infection, and before the decision regarding hospital discharge was determined.

### 2.7 Statistical analysis

All analyses were performed using Stata version 16, and *p*<0.05 was considered statistically significant. Data were described as the mean and standard deviation or frequency (n and %) and compared between groups using Student’s t-test or the chi-square and Fisher’s exact test for continuous and categorical data, respectively. A multivariate analysis was performed to adjust for potential confounding effects on the risk of pneumonia in the AP treatment and control groups. The treatment effect model was applied after inverse probability-weighted regression adjustment. First, a treatment model was constructed using a logit model by fitting the treatment to the covariates (i.e., age 51-60 years, body mass index >25-35 kg/m^2^, and clinical symptoms, including fever, cough, nasal congestion or runny nose, myalgia, loss of smell, loss of taste, and diarrhoea). Second, the outcome model was constructed using Poisson regression by fitting the pneumonia outcome to the covariates. The risk of pneumonia, risk difference, and number needed to treat were estimated.

## 3. Results

A total of 528 patients were included. There were 243 patients in the AP treatment group and 285 patients in the control group (Table 1). The two study groups were comparable in terms of average age, sex, and body mass index; however, the age group distribution was slightly different (*p*=0.035). All 243 patients in the AP treatment group and 53.0% (151/285) in the control group were symptomatic at the time of hospital admission. The proportion of patients with each symptom, except for diarrhoea, was higher in the AP treatment group than that in the control group. In the AP treatment group, 88.1% (214/243) received AP extract and 11.9% (29/243) received crude AP. The AP treatment was started within a week after onset of symptoms in 93.8% of the patients, i.e., 0-3 days, 53.1% (129/243); 4-7 days, 40.7% (99/243); and 8-13 days, 6.2% (15/243). In the control group, approximately half of the patients were asymptomatic. The risk of pneumonia in this group was similar for patients with asymptomatic and mild COVID-19 (*p*=0.340), and it progressively increased with older age (*p*=0.001) (Table 2).

**Table 1.** Baseline characteristics of COVID-19 patients in the *Andrographis paniculata* (AP) treatment and control groups.

| Characteristics | AP-treatment<br>(n=243) | Control<br>(n=285) | *p*-value |
| --- | --- | --- | --- |
| Age (years), mean ± SD | 37.7 ± 11.1 | 37.0 ± 11.2 | 0.528 |
| Age group (years), n (%) |  |  |  |
| 15-30 years* | 84 (34.6) | 79 (27.7) | 0.035 |
| 31-40 years | 69 (28.4) | 80 (28.1) |  |
| 41-50 years | 51 (21.0) | 90 (31.6) |  |
| 51-60 years | 39 (16.0) | 36 (12.6) |  |
| Male, n (%) | 94 (38.7) | 106 (37.2) | 0.725 |
| Body mass index (kg/m<sup>2</sup>), mean ± SD | 23.7 ± 4.1 | 24.0 ± 4.2 | 0.544 |
| Being symptomatic on admission, n (%) | 243 (100.0) | 151 (53.0) | <0.001 |
| Presenting symptoms, n (%) |  |  |  |
| Fever | 85 (35.0) | 43 (15.1) | < 0.001 |
| Cough | 136 (56.0) | 80 (28.1) | < 0.001 |
| Nasal congestion or runny nose | 69 (28.4) | 51 (17.9) | 0.004 |
| Myalgia | 37 (15.2) | 16 (5.6) | < 0.001 |
| Loss of smell | 81 (33.3) | 49 (17.2) | < 0.001 |
| Loss of taste | 30 (12.3) | 12 (4.2) | 0.001 |
| Diarrhea | 10 (4.1) | 13 (4.6) | 0.802 |

\*Ten patients in the AP-treatment and 23 patients in the control group were adolescent (15 to 20 years).

**Table 2.**
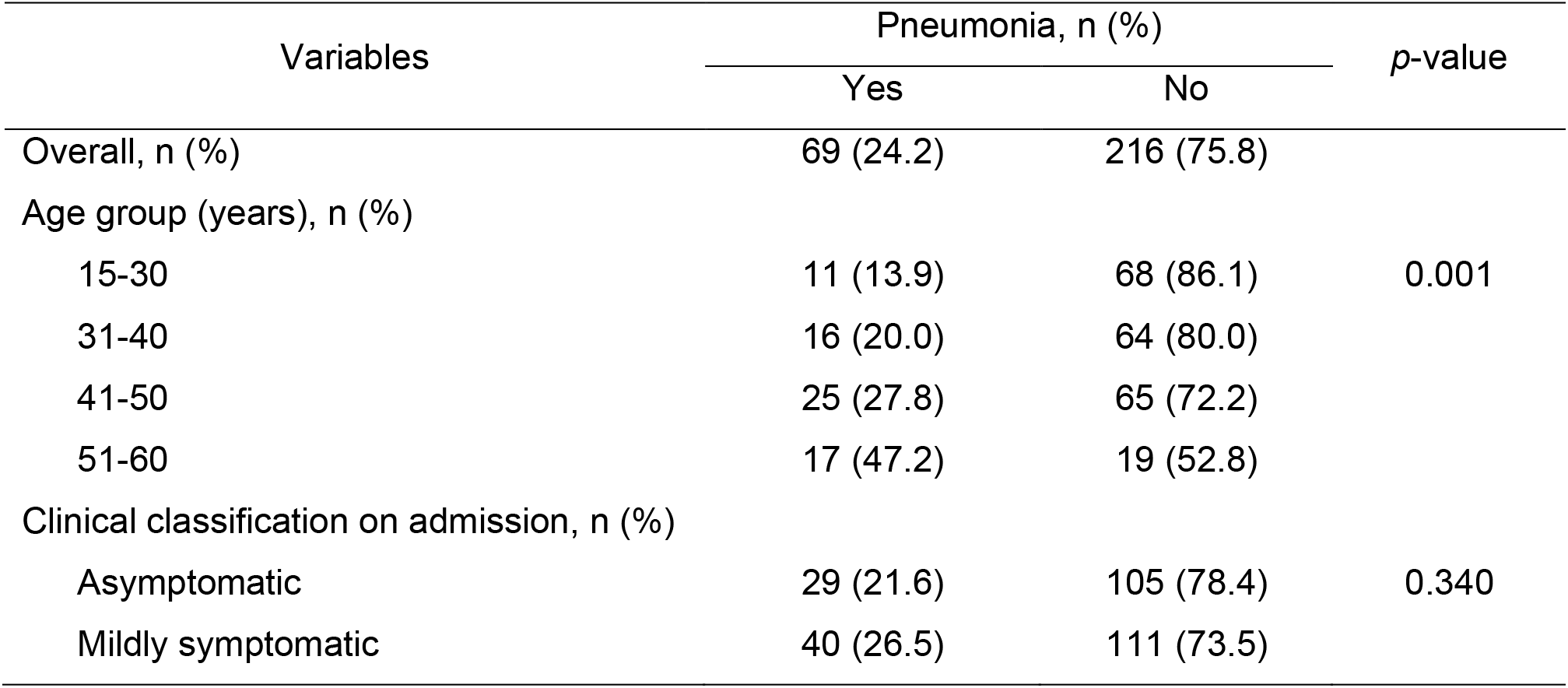
Risks of pneumonia by subgroups in 285 early-stage (asymptomatic or mildly symptomatic) COVID-19 patients in the control group without *Andrographis paniculata* treatment.

The unadjusted risk of pneumonia for each potential risk variable was explored using a univariate analysis (Table 3). Pneumonia occurred in one patient in the AP treatment group and 69 patients in the control group. The AP treatment patient who developed pneumonia was a 36-year-old man who received the crude AP regimen at 11 days after symptom onset. The adjusted risk of pneumonia in the control group was 24.3% (95% confidence interval [CI], 19.0%-29.7%); however, it was 0.3% (95% CI, 0%-0.9%) in the AP treatment group, resulting in a risk difference of 24.0% (95% CI, 18.7%-29.4%) (Table 4).

**Table 3.** The unadjusted risk of pneumonia for each potential risk variable in 528 early-stage (asymptomatic or mildly symptomatic) COVID-19 patients: A univariate analysis.

| Potential risk variables | Pneumonia, n (%) |  | p-value |
| --- | --- | --- | --- |
|  | Yes | No |  |
| AP-treatment*, n (%) |  |  |  |
| Yes | 1 (0.4) | 242 (99.6) | <0.001 |
| No | 69 (24.2) | 216 (75.8) |  |
| Age group, n (%) |  |  |  |
| ≤ 50 years | 53 (11.7) | 400 (88.3) | 0.009 |
| 51-60 years | 17 (22.7) | 58 (77.3) |  |
| Sex |  |  |  |
| Male | 26 (13.0) | 174 (87.0) | 0.892 |
| Female | 44 (13.4) | 284 (86.6) |  |
| Body mass index categories, n (%) |  |  |  |
| ≤ 25 kg/m<sup>2</sup> (thin or normal) | 42 (11.8) | 315 (88.1) | 0.144 |
| >25-35 kg/m<sup>2</sup> (overweight or obese) | 28 (16.4) | 143 (83.6) |  |
| Presenting symptoms, n (%) |  |  |  |
| -History of fever |  |  |  |
| Yes | 13 (10.2) | 115 (89.8) | 0.235 |
| No | 57 (14.2) | 343 (85.8) |  |
| -High body temperature (>37.5°C) |  |  |  |
| Yes | 3 (18.8) | 13 (81.3) | 0.511 |
| No | 67 (13.1) | 445 (86.9) |  |
| -Cough |  |  |  |
| Yes | 23 (10.6) | 193 (89.4) | 0.141 |
| No | 47 (15.1) | 265 (84.9) |  |
| -Nasal congestion or runny nose |  |  |  |
| Yes | 12 (10.0) | 108 (90.0) | 0.231 |
| No | 58 (14.2) | 350 (85.8) |  |
| -Myalgia |  |  |  |
| Yes | 4 (7.5) | 49 (92.5) | 0.196 |
| No | 66 (13.9) | 409 (86.1) |  |
| -Loss of smell |  |  |  |
| Yes | 12 (9.2) | 118 (90.8) | 0.119 |
| No | 58 (14.6) | 340 (85.4) |  |
| -Loss of taste |  |  |  |
| Yes | 2 (4.8) | 40 (95.2) | 0.091 |
| No | 68 (14.0) | 418 (86.0) |  |
| -Diarrhea |  |  |  |
| Yes | 3 (13.0) | 20 (87.0) | 0.975 |
| No | 67 (13.3) | 438 (86.7) |  |

\*AP = *Andrographis paniculata*

**Table 4.** Effect of *Andrographis paniculata* (AP) treatment on pneumonia development in adolescents and adults with early-stage (asymptomatic or mildly symptomatic) COVID-19, after adjustment for potential confounding effects

|  | Point estimate | (95% CI) |
| --- | --- | --- |
| Risks of pneumonia by AP-treatment, % |  |  |
| No | 24.3 | (19.0, 29.7) |
| Yes | 0.3 | (0.0, 0.9) |
| Risk difference, % | 24.0 | (18.7, 29.4) |
| Number needed to treat (NNT), n | 4 | (3, 5) |
| Number prevented when 100 patients are treated, n | 24 | (19, 29) |

NNT = 1 / risk difference; the number of patients that needed to treat for one patient not to have pneumonia

Therefore, the number needed to treat to avoid pneumonia development in one patient with early-stage COVID-19 was four (95% CI, 3-5). Treating 100 early-stage COVID-19 patients with the AP regimen prevented 24 (95% CI, 19-29) cases of pneumonia. Abnormal symptoms suggesting adverse effects of AP treatment were detected in eight patients (nausea, 3; diarrhoea, 2; dizziness, 1; tachycardia, 1; chest tightness, 1). No substantial or persistent adverse effects of the AP treatment were observed in the study population.

## 4. Discussion

Our study demonstrated that the AP regimens orally administered within one week of the onset of symptoms were effective and safe for the treatment of early-stage COVID-19 and prevention of disease progression to moderate COVID-19 with pneumonia. These findings are in agreement with those of a previous study that reported a small, randomised, controlled trial involving adults with mild COVID-19 (Wanaratna et al., 2022). Furthermore, a study performed in China demonstrated that a semi-synthetic injectable andrographolide preparation (Xiyanping) effectively improved the recovery of patients with mild-to-moderate COVID-19 (Zhang et al., 2021). All patients in the AP treatment group had mild symptoms when AP treatment was initiated. This was not unexpected because of the shortage of AP products and because AP is a well-known treatment for acute respiratory tract infections (Coon, 2004; Poolsup, 2004). Almost 90% of the AP treatment patients in this study were treated with the AP extract regimen; therefore, we recommend the AP extract regimen (60 mg of andrographolide per dose, 3 times daily for 5 days). Further studies of the therapeutic effects of the crude AP regimen are necessary. AP treatment should be started as soon as possible during early-stage COVID-19 to effectively combat viral replication and inflammatory reactions that cause more severe diseases.

There are a few possible explanations for the reduced risk of pneumonia during early-stage COVID-19 after AP treatment. First, andrographolide can inhibit intracellular SARS-CoV-2 replication (Phumiamorn et al., 2020; Sa-Ngiamsuntorn et al., 2021). Second, its anti-inflammatory effects are related to the reduced expression of interleukin-6 and tumour necrosis factor-alpha (Zhu et al., 2021) which cause tissue damage. Third, it systemically distributes to various organs, including the lungs (Songvut et al., 2022). It was found that C-reactive protein, an inflammatory biomarker that is stimulated by interleukin-6, is related to the severity of COVID-19 (Mueller et al., 2020). A randomized, controlled trial of AP extract treatment for 63 adults with mild COVID-19 suggested its promising efficacy for preventing pneumonia, shortening of the viral shedding period, and suppressing C-reactive protein (Wanaratna et al., 2022). Tocilizumab, which is an anti-interleukin-6 monoclonal antibody that reduces C-reactive protein production, has been recommended as an effective treatment for severe COVID-19 (Xu X, et al., 2020; World Health Organisation, 2022b). The anti-inflammatory and immunomodulatory properties of AP (Hossain et al., 2021; Zhu et al., 2021; Zeng et al., 2022) make it a potential treatment option that can decrease inflammation and relieve COVID-19 symptoms; furthermore, it has specific anti-SARS-CoV-2 effects.

The AP treatment regimens in this study resulted in a few cases of mild gastrointestinal discomfort and non-specific mild symptoms; however, substantial or persistent adverse drug reactions were not observed. In addition, information regarding the acceptable safety of AP was determined by evaluating the long history of herbal medicine practices and recent reviews (Dai et al., 2019; Hossain et al., 2021; Worakunphanich et al., 2021; Zeng et al., 2022). During animal studies, it was found that andrographolide has organ-protective effects on the liver, kidney, lung, and gastrointestinal tract. Toxicity is related to the concentration and administration time of andrographolide (long-term, high-dose, or rapid high-dose administration). Some individuals experience allergic reactions ranging from minor skin rashes to serious anaphylaxis when exposed to AP; however, these reactions are generally observed with high doses. Reproductive toxicity and nephrotoxicity observed during animal studies occurred when much higher doses or semi-synthetic andrographolide were used (Zeng et al., 2022).

In the control group, the risk of pneumonia for asymptomatic patients was similar to mildly symptomatic patients. It is well-established that some asymptomatic patients are in pre-symptomatic phase and later develop symptoms (National Institutes of Health USA, 2021). Approximately 20% of patients in the control group who were asymptomatic at the time of admission developed pneumonia. The community-wide active case finding in the included provinces might be responsible for the very early detection of pre-symptomatic COVID-19 patients. Further study on AP treatment for asymptomatic COVID-19 patients is needed. However, it is likely that AP treatment could prevent pneumonia in early-diagnosed asymptomatic patients.

The disadvantages of a retrospective cohort study are that the quality of the data is generally inferior to that of a prospective study, selection bias may occur, and the presence of confounders may distort the association of the study variables. During this study, the structured medical records of COVID-19 patients were prepared for prospective recording. We randomly and blindly selected the control group to prevent selection bias. Additionally, the AP treatment and control groups were from the same population and matched by inclusion criteria, and the existing potentially confounding effects were statistically adjusted.

Regarding generalizability, we explored the characteristics of the control group and compared them with those of patients involved in other COVID-19 studies published in the literature. The finding that nearly half of the patients in our control group were asymptomatic at the time of admission is in agreement with the findings of another study performed in a similar setting in Chennai, India (Krishnasamy et al., 2021). In our control group the occurrence of pneumonia was approximately 24%, which is comparable to the 20% occurrence rate of moderate and severe COVID-19 reported by a study of a large population in China (Wu and McGoogan, 2020). Additionally, the progressively increasing risk of pneumonia for older age groups is similar to that observed in the United States of America surveillance data regarding the risk of COVID-19 hospitalisation (Centers for Disease Control and Prevention USA, 2022a). During the study period, approximately 90% of the circulating strains in the included provinces belonged to the B.1.36.16 lineage (COVID-19 Network Investigations Alliance, 2021), and the COVID-19 vaccine was not yet available.

Oral AP treatment comprising the recommended andrographolide dosage is ideal for early-stage COVID-19 which is generally self-limited but could progress. Furthermore, this oral medicine that occurs in nature is effective, safe, economical, and widely available. This treatment can result in the prevention of more severe COVID-19 and decreased hospitalisation and treatment costs. Additional information is needed to explore its effects on children, patients older than 60 years, those with underlying diseases, and pregnant women. Because AP is an herbal product, quality control and the standardisation of its preparations are important issues (Gagnier et al, for the CONSORT Group, 2006).

In conclusion, in addition to general supportive care, the AP treatment regimens administered to adolescent and adult patients with early-stage COVID-19 and without comorbidities or pregnancy are safe and effective for preventing disease progression to pneumonia. The treatment effects of andrographolide may vary with different circulating SARS-CoV-2 strains and herb immunity.

## Data Availability

All data produced in the present study are available upon reasonable request to the authors.

## Abbreviations

AP: Andrographis paniculata
COVID-19: coronavirus disease 2019
SARS-CoV-2: severe acute respiratory syndrome coronavirus 2

## Acknowledgement

We thank the involved personnel at the study hospitals in Samut Sakorn, Nakhon Pathom, and Ratchaburi provinces, for their assistance in data collection.

## CRediT authorship contribution statement

AB: Conceptualisation, Project administration, Review, Funding acquisition, Editing & Resource; TS: Methodology, Review, Editing & Resource; TT: Methodology, Review, Editing & Resource; KW: Visualisation, Methodology, Review & Editing; PC: Formal analysis, Manuscript drafting, Review & Editing; CR: Formal analysis; SB: Formal analysis; SC: Formal analysis; SB: Formal analysis, Review & Editing; SL: Formal analysis, Visualisation, Review, Manuscript drafting, Editing & Funding acquisition; SS: Conceptualisation, Formal analysis, Visualisation, Manuscript drafting, Review & Editing.

## Ethical approval

The study was approved by the Department of Thai Traditional and Alternative Medicine, Ministry of Public Health, Thailand. Ethical approval was obtained from the Human Research Ethics Committee of Siam University (Reference Number: 2021/005). The data presented here were recorded during routine clinical practice. All data were de-identified prior to analysis and all the authors had all necessary administrative permissions to access the data.

## Consent for publication

Not applicable

## Highlights

- The *Andrographis paniculata* treatment in early-stage COVID-19 prevent pneumonia.
- *Andrographis paniculata* dosage regimens for COVID-19 are acceptably safe.
- Andrographolide inhibits virus replication, so the treatment should be started early.
- To prevent one early-stage COVID-19 patient from pneumonia, four need to be treated.
- The *Andrographis paniculata* treatment is effective, safe, and economical for COVID-19.

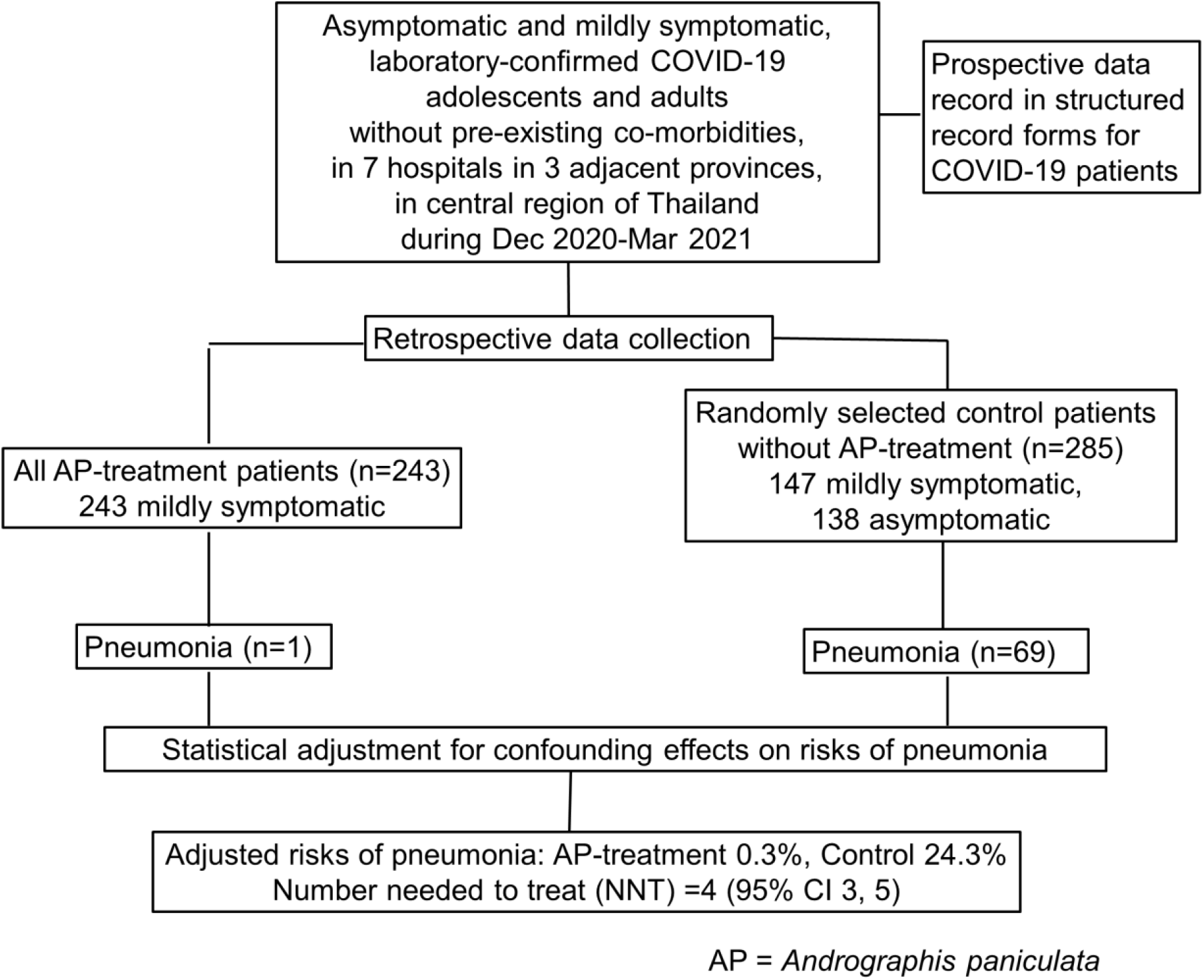

